# Efficacy of Famotidine for COVID-19: A Systematic Review and Meta-analysis

**DOI:** 10.1101/2020.09.28.20203463

**Authors:** Rahul Sethia, Manya Prasad, Soumya Jagannath Mahapatra, Neeraj Nischal, Manish Soneja, Pramod Garg, Shalimar

## Abstract

**Background:** Coronavirus Disease 2019 (COVID-19) pandemic continues unabated in many parts of the world. In the absence of any definite antiviral therapy except some benefit of remdesivir, there is an ongoing search for effective therapy. Famotidine has been shown to reduce mortality in hospitalized patients in a few studies. We conducted a systematic review on the use of famotidine in COVID-19.

**Methods:** We searched the databases Medline, Embase, Cochrane CENTRAL and Medrxiv. Title/abstract screening, full text screening and data abstraction were carried out in by two reviewers. Case series, cohort studies and randomized trials were included.

**Results:** Five studies were eligible for inclusion: all were retrospective cohort or case series. Low quality evidence suggests a likely clinical benefit for the use of famotidine in decreasing mortality in hospitalized patients with moderate to severe COVID-19. A meta-analysis of two cohort studies showed a statistically significant decrease in the composite outcome for death and intubation with famotidine (HR 0.44, 95% CI 0.27 to 0.73).

**Conclusion:** Further evidence from RCTs is required for famotidine to treat COVID 19.

## 1. Introduction

Coronavirus Disease 2019 (COVID-19), the illness caused by Severe Acute Respiratory Syndrome Coronavirus 2 (SARS-CoV-2) continues to pose enormous burden on healthcare system with significant morbidity and mortality. As of 19th September 2020, COVID-19 has resulted in close to one million deaths worldwide.(1)

There is no proven effective therapy for COVID-19. Remdesivir is the only antiviral that has shown some clinical benefit but without any reduction in mortality. A globally implemented, safe vaccination program seems to be the only hopeful long term solution, but is at least 6 months away. Moreover, the safety and efficacy of any vaccine candidate is yet to be proven. Therefore, there is a need for effective therapy to reduce mortality.

Famotidine, a H2 receptor antagonist, has been in clinical use for many decades for reducing gastric acid production in the treatment of peptic ulcer and gastroesophageal reflux disease. It has an excellent safety profile and being off patent, is cheap. Famotidine has been shown to bind papain-like protease (PLpro) and 3 chymotrypsin-like protease (Mpro) of SARSCoV-2 in *in silico* molecular docking studies.(2) However, the inhibition has not been confirmed. Famotidine has been shown to reduce mortality in hospitalized patients with COVID-19 in a few studies.(3–5) We conducted a systematic review to assess the effect of famotidine on the clinical outcomes in patients with COVID-19.

### Objective

To assess the effectiveness and safety of famotidine in patients with COVID-19.

## 2. Methodology

### 2.1. Methods

The Preferred Reporting Items for Systematic Reviews and Meta-analyses (PRISMA) was adhered to in the present report.(6)

### 2.2. Inclusion criteria

The following inclusion criteria were used for eligibility:

Type of participants: We included studies on patients with all grades of severity of COVID-19.

Type of interventions: We included studies assessing famotidine in humans with COVID-19.

Type of outcomes: We included studies reporting the following outcomes:

Primary outcome: Overall mortality

Secondary outcomes:

- Clinical recovery (as defined by authors)
- Rate of ICU admission
- Length of ICU stay
- Length of hospital stay
- Need for mechanical ventilation
- Viral clearance
- Adverse events

Type of studies: We included randomized controlled trials, cohort studies and case series.

### 2.3. Data sources and searches

We searched the following databases for articles published till 20^th^ September 2020: Medline, Cochrane CENTRAL, Trip database, Pubmed (for articles not yet indexed in Medline) and Medrxiv and SSRN for pre-print articles.

Editorials, letters, news, reviews, expert opinions, case reports, and studies without original data were excluded.

Reference lists of retrieved articles and pertinent reviews were also searched for relevant articles. No language restriction was imposed.

### 2.4. Selection of studies

Titles and abstracts were screened by two reviewers (RS, PG) independently. Full texts of articles considered potentially eligible were obtained. The data were abstracted by two reviewers independently and risk of bias was assessed. Disagreement was resolved by discussion.

### 2.5. Data extraction

The following data were extracted: study design, inclusion criteria, number of patients, patient characteristics, dose, duration and timing of famotidine, co-medications, outcomes and method of adjustment (for cohort studies and case series).

### 2.6. Risk of Bias assessment

Risk of bias was assessed using the revised version of Newcastle Ottawa Scale for Cohort studies and the JBL checklist for case series. (7,8)

### 2.7. Data synthesis and statistical analysis

We calculated pooled risk ratio and 95% CI using the Generic Inverse Variance approach. Random effects model was used to conduct the meta-analysis. We carried out the statistical analysis using Review Manager 5.3. Heterogeneity was assessed using visual inspection of forest plot and the I^2^ statistic.

### 2.8. GRADE

We used GRADE methodology to rate certainty of evidence for outcomes as high, moderate, low or very low (9).

## 3. Results

### 3.1. Study selection

Our search yielded 13 titles and abstracts - all were identified from the electronic database search. We excluded 8 articles based on a review of the title and abstract, leaving 5 articles for full review. These five studies were found eligible on full text screening; one of these was a pre-print articles that has not been peer reviewed. These 5 studies were included in the systematic review. (4,5,3,10,11). (Fig 1)

**Figure 1:**
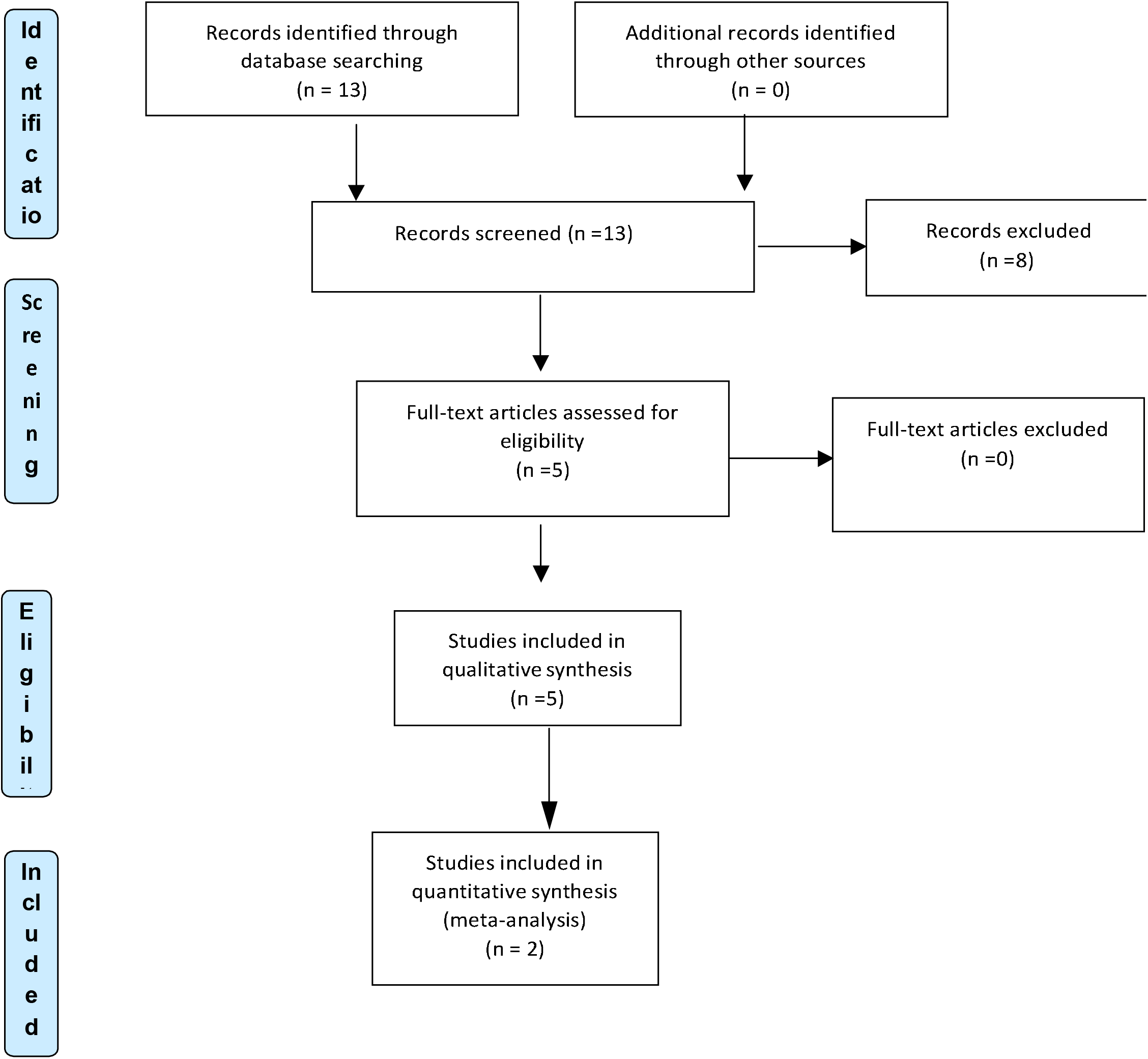
PRISMA flow diagram for study selection

### 3.2. Study characteristics and estimates reported

The studies included were 3 cohort studies and 2 case series. All studies included patients hospitalized with COVID-19. Three cohort studies (3,4,11) and one case series (10) included mostly patients with severe COVID-19, and one case series included patients with mild to moderate COVID-19.(5)

#### Efficacy of Famotidine: Descriptive Analysis (Table 1)

The five selected studies included a total of 2643 patients with COVID-19 of whom 312 patients received famotidine. All except 10 patients were hospitalized with moderate-severe illness. The dose of famotidine varied from 40-233 mg/day given for 5-21 days. Two cohort studies including 84 and 83 patients who received famotidine with contemporary controls – 1536 and 795 patients who did not receive famotidine respectively – showed a reduction in mortality: 58% (HR 0.42 95 % CI 0.21-0.85, p=0.02) and 63% (OR 0.37, 95% CI 0.16-0.86, p=0.02) (5,7). One study that compared mortality with published series of patients with similar severity showed mortality reduction of around 26-45% with the use of combined famotidine and cetrizine.(11) One case series of 25 hospitalized patients with moderate disease showed zero mortality.(10) Another case series of 10 non-hospitalized patients reported improvement in patient reported outcomes.(5)

**Table 1:** Summary results of the included cohort studies.

| S. no. | Author | Type of study | Patients: number, age | Intervention (Famotidine) Dose, duration (n) | Controls (no Famotidine) (n) | Severity of illness | Outcome | Overall benefit |
| --- | --- | --- | --- | --- | --- | --- | --- | --- |
| 1. | Freedberg et al. (4) | Retrospective cohort | 1620 hospitalized patients | 84 patients (5.1%)<br>Median dose 136 mg (63-233 mg)<br>Median duration 5.8 days | 1536<br>Propensity matched | COVID-19 hospitalized patients with moderate – severe illness but not intubated | Death or intubation In 8/84 (10%) of Famotidine vs. 332/1536 (22%) non-Famotidine (HR 0.42 95 % CI 0.21 -0.85) | Yes |
| 2. | Janowitz et al. (5) | Case series | 10 patients with non - hospitalized COVID-19 | 10 patients<br>Dose ( 20- 80 mg thrice daily)<br>Median duration 11 (5-21) days | None | Non-hospitalized patients | Improvement in patient reported outcomes | Yes |
| 3. | Mather et al. (3) | Retrospective cohort | 878 hospitalized patients<br>Mean age (67 +/- 16 years) | 83 patients (9.5%)<br>Dose (20-160 mg/day)<br>Median duration 4 (2-8) days | 795 | 239 patients (27.2%) required mechanical ventilation, 191 patients (21.8%) died | Famotidine use associated with decrease risk of in-hospital mortality (OR 0.37, 95% CI 0.16-0.86, p =0.02), decreased combined death or intubation: 30 /83 vs. 400/795(OR 0.47, 95% CI 0.23-0.96, p =0.04) | Yes |
| 4. | Tomera et al (10) | Case series | 25 hospitalized patients<br>Median age 62 (32-80) years | 25 patients treated with famotidine – 40 mg bd plus celecoxib (400 mg bid) | None | 17/25 (68%) required oxygen therapy on admission<br>9/25 (36%) had extremely high LDH (>365) with Wuhan model prediction of >98% mortality | No mortality<br>No need of mechanical intubation/renal replacement therapy<br>Median hospital stay 3 (1-16) days | Yes |
| 5. | Hogan et al (11) | Retrospective Cohort | 110 hospitalized patients, mean age 63.7+/- 18.1 years | 110 patients given famotidine - 20 mg bid plus cetirizine 10 mb bid | Published reports of Covid- 19 inpatients of similar severity | Severe or critical disease (intubation rate-16,4%<br>Mean comorbidities – 2.7 per patient.<br>Only 30% given steroids, 50% given Tocilizumab | Combination therapy resulted in mortality reduction of 26-45% compared to published reports of similar patients | Yes |

### 3.3. Risk of Bias assessment

In all three cohort studies, risk of bias was low for selection of exposed and non-exposed population and assessment of exposure (3, 4, 11). All three cohort studies were assessed to have low risk of bias from outcome being present at the start of the study. However, adequate adjustment and assessment of prognostic factors were not carried out by one study (11). Follow up was adequate for all outcomes in the cohort studies; however, all cohort studies were assessed as being high risk of bias for co-interventions being dissimilar in the two groups.

Both case series reported consecutive and complete inclusion of participants and the condition being measured in a standard, reliable way (5, 10). Both also clearly reported the demographics of participants and clinical information. However, one case series did not report the criteria for inclusion.

### 3.4. Pooled effect of Famotidine on composite endpoint of death and intubation

A meta-analysis of two cohort studies showed a statistically significant decrease in the composite outcome for death and intubation with the use of Famotidine (HR 0.44, 95% CI 0.27 to 0.73). There was no heterogeneity in this meta-analysis (I^2^=0%) (Fig 2)

**Figure 2:**
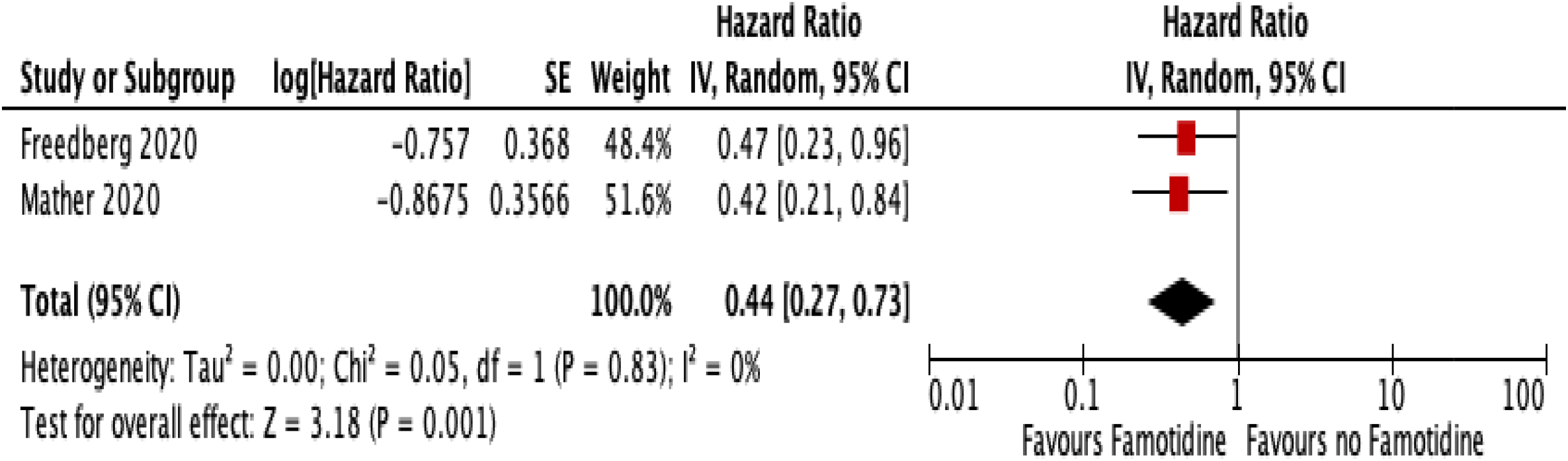
Effect of Famotidine on composite endpoint of death and intubation in COVID-19

### 3.5. Certainty in Evidence

The certainty in evidence was assessed as very low using the GRADE approach. The certainty was rated down for risk of bias, as the included studies were observational studies with possible residual confounding. (Table 2)

**Table 2:** Critical analysis of included cohort studies.

| S no. | Author | Merits of study | Demerits of study |
| --- | --- | --- | --- |
| 1 | Freedberg et al.(4) | 1. First study to show decrease in death / rate of intubation in patients with covid-19.<br>2. Propensity matched score to eliminate bias.<br>3. Speculated MOA of famotidine by decreasing cytokine storm looking into Hs CRP, serum ferritin. | 1. Not an RCT. Retrospective observational single center study.<br>2. Variable dose of famotidine and its indication prior to hospitalisation not clear. Poor documentation - SOC not mentioned.<br>3. Formulation of famotidine used not mentioned. |
| 2 | Janowitz et al.(5) | 1. Preliminary study. Proof of concept. | 2. Case series. Not an RCT.<br>3. All patient recovered and reported improvement , patient may recover due to natural course of disease – placebo effect .<br>4. Recall bias – based on telephonic assessment of symptoms. |
| 3 | Mather et al.(3) | 1. Propensity matched score to eliminate bias.<br>2. Use of famotidine associated with decrease in markers of covid -19 disease severity such as CRP, calcitonin etc.<br>3. Changes in hs CRP and lymphocyte: neutrophil ratio demonstrates the mechanistic insight about the efficacy of famotidine.<br>4. Selection of patients with multiple comorbidities. | 1. Retrospective single center observational study.<br>2. Variable dose of famotidine.<br>3. SOC – 50 % patients were given steroids/HCQ/azithromycin – confounding effect in improvement.<br>4. Despite propensity score matching, there was a difference in age of patients which nearly approached statistical significance, (famotidine being younger). Age is the single most important determinant of outcome in COVID. Therefore it is possible that Type-2 error may have played a role in this study and the findings needs confirmation in a larger sample size. |
| 4 | Tomera et al.(10) | 1. 100 % survival with improved radiological outcome as well as statistically significant improvement in clinical and biochemical parameters.<br>2. No drug related adverse event. No report of increased AKI with celecoxib use.<br>3. High risk patients – BMI > 30. All patients discharged , off oxygen<br>4. COX-2 inhibition and use of antihistaminics - potential target to decrease inflammatory cascade. | 1. Case series. No control arm.<br>2. Small number of patients , use of remdesvir in some patients and not in some patients not clear.<br>3. Effect of concomitant antiviral therapy – hard to discern in small case series. |
| 5 | Hogan et al.(11) | 1. Proof of concept study<br>2. Demonstrated decline in severity of pulmonary symptoms, increased discharge rate in severe patients. | 1. No comparative analysis between survivors and non-survivors. To be done in view of multiple confounders.<br>2. No details of assessment of severity of COVID in patients.<br>3. Comparative analysis with SOC group of other countries was to be done. |

**Table 3.**
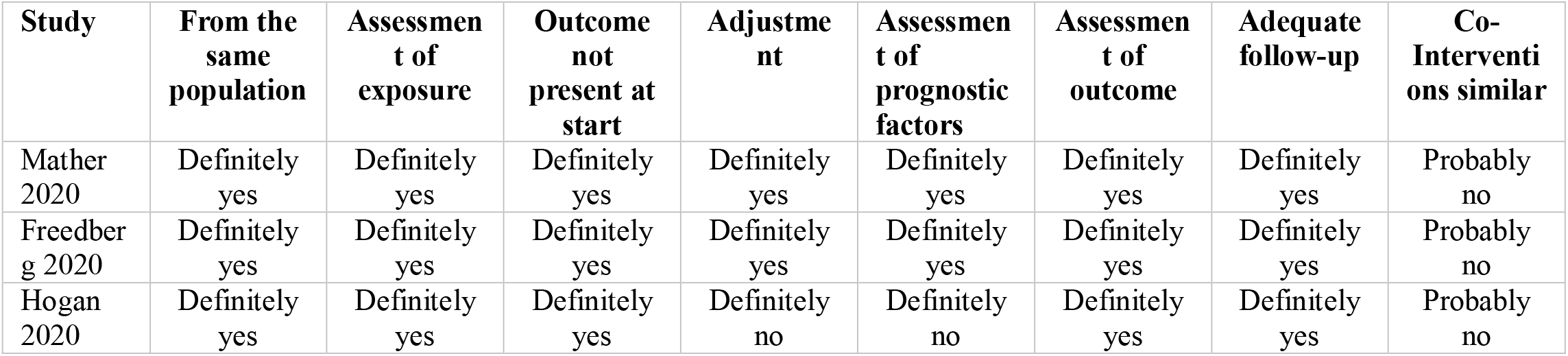
Risk of bias of included cohort studies.

**Table 4.**
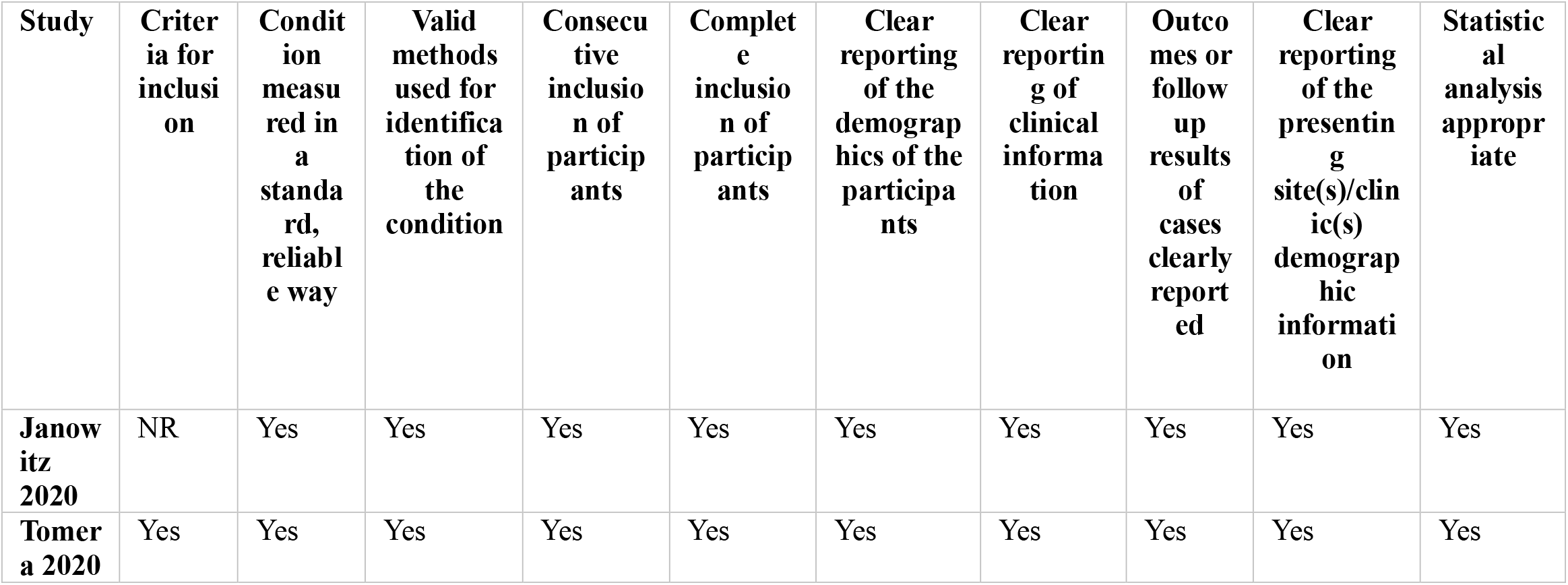
Risk of bias of included case-series.

**Table 5.**
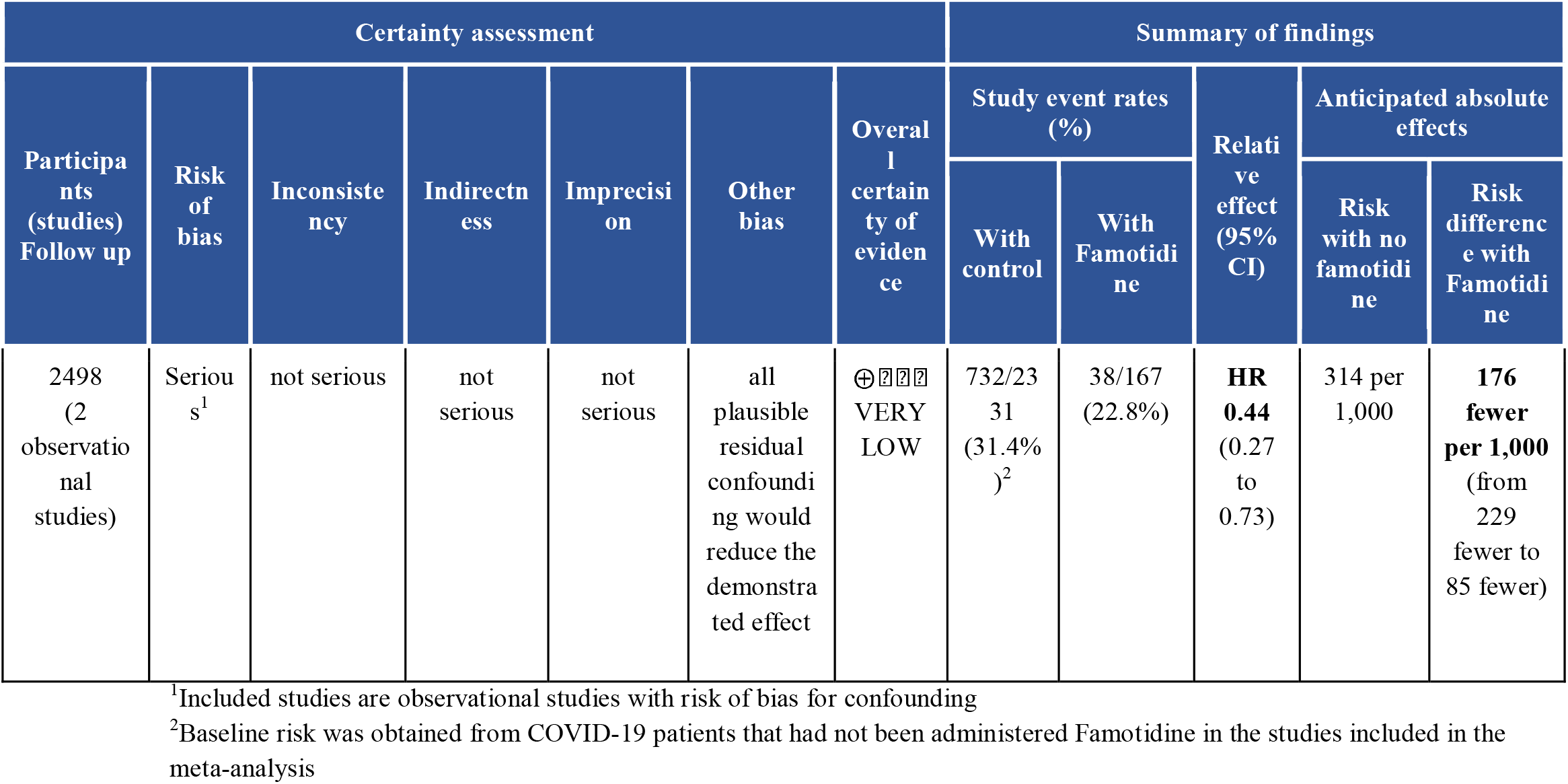
Summary of findings and GRADE profile for the composite of death and intubation

## 4. Discussion

The present systematic review found that the use of famotidine was associated with a reduction in the composite endpoint of death and the requirement of mechanical intubation in patients with COVID-19. The included studies were retrospective, heterogeneous, with a small sample size and a variable dose and route of administration of famotidine.

The available data suggest 56% reduction in mortality and requirement of mechanical intubation among patients treated with famotidine and standard of care (SOC) as compared to those managed with SOC alone. Cytokine storm plays an important role in the clinical severity and outcome of COVID-19. However, the exact mechanism of action of famotidine, a H2 receptor antagonist (H2RA) in modulating the clinical outcome among COVID-19 patients is unclear. Multiple theoretical hypotheses have been proposed for the effect of famotidine. Pathophysiologically, histamine plays a role in the clinical symptomatology of COVID-19 including sore throat, cough, diarrhea, myalgia, anosmia, and skin rashes. The available literature suggests that H2RAs have a modulatory effect on both innate (neutrophils, monocytes, macrophages, dendritic cells, and natural killer cells) and adaptive (CD4+, CD8+, T-reg and B cells) immune system.(12,13). Famotidine has been shown to bind papain-like protease (PLpro) and 3 chymotrypsin-like protease (Mpro) of SARS-CoV-2 in *in silico* molecular docking studies.(2) The mechanism of action specific to SARS-CoV2 appears to be distinct from the acid suppression mechanism as a recent Korean study reported worse outcomes in patients with active proton pump inhibitor usage.(14)

Many important clinical issues such as ideal dose, duration, timing of initiating the treatment after the onset of symptoms, and route of administration of famotidine, which may be associated with an improvement in the clinical outcome are unclear. The available studies included variable doses, duration of therapy, and route of administration of famotidine. Freedberg et al. reported dose varying from 10 mg-40 mg over a median duration of 5.8 days with a cumulative dose of 136 (63-233) mg. In the study by Marther et al, oral administration was reported in 83% of cases with 20 mg/d in 95% and remaining received 40 mg/d. The cumulative dose was 80 mg (40-160 mg) given over 4 days. Janowitz et al. used a dose ranging from 60 mg/d to 240 mg/day over a median duration of 11 days (range 5-21 days). An ongoing multicentre RCT (NCT0437020262) comparing standard of care (SOC) with SOC plus famotidine in the US is administering intravenous famotidine at a dose of 360 mg/d for 14 days.(15)

The present systematic review and meta-analysis is the first, to the best of our knowledge, to address the role of famotidine in COVID-19. It incorporates a comprehensive search of three major databases, as well as pre-print articles. In addition, we used to GRADE approach to rate the certainty in evidence, thus paying due attention to methodological issues like risk of bias, imprecision, indirectness and inconsistency.

There are many limitations of the present study which are inherent to the included studies. First, the studies included in the analysis had a small number of patients treated with famotidine. Studies were single-center and retrospective observational studies with their associated bias.

Multiple other drugs were used for the management of patients, which could have influenced the clinical outcomes

In conclusion, the available evidence suggests a potential role of famotidine in the management of COVID-19, which needs to be explored in randomized controlled trials.

## Data Availability

None

## Funding

No funding received

## Conflicts of interest

None declared

## Notes

### Competing Interest Statement

The authors have declared no competing interest.

